# Seasonal Variations and the Associated Factors of Acute Appendicitis at a Tertiary Hospital: A Case of SPHMMC, Addis Ababa, Ethiopia

**DOI:** 10.1101/2024.04.06.24305309

**Authors:** Lohide Daniel, Fekadu Negash

## Abstract

**Background:** Acute appendicitis, a surgical emergency, is a prevalent pathology with uncertain etiology, influenced by various factors and seasonal variations.

**Objective:** To determine the seasonal variations and the associated factors of Acute Appendicitis.

**Methods:** The study analyzed demographic features, seasonal variation, length of hospital stay, and surgical treatment outcomes in 384 patients treated from 01/01/2022 to 31/12/ 2022 G.C. The patients were presented in four different seasons; December-February as winter, March-May as autumn, June-August as summer, and September-Nov as spring, which were assessed to describe seasonal variation and compare to what are observed in other countries. SPSS version 26 was used for all the statistical assessments and analyses.

**Results:** Out of all 384 patients, 64.3% were males and 35.7% were females. The mean age was 20.8, male: female, 1.8:1. The majority of patients were in their 2nd and 3rd decades of life, and more common in the male 10–19 age group (22.1%). 23.9% of cases were observed in autumn, 22.7% in winter, 20.6% in spring, and 32.8% in summer. 10.1% increase in summer as compared to winter, 8.9% increase as compared to autumn, and 12.2% increase as compared to spring. The highest admissions were in the summer (32.8%). July had the highest number of cases (12.5%), which coincided with heavy rainy months, the rainiest days, and the lowest sunlight hours. The highest rainfall was observed in July (386mm) and August (491 mm), and the lowest sunlight hours were observed in July (6.1 hours) and August (5.9 hours), respectively. Monthly cases correlated positively with rainfall (Pearson’s R =.637, p<.05) and negatively with sunlight hours (Pearson’s R = -.618, p<.05). Complicated cases were slightly more common (50.9%), more in the 10–19 age group. Males had more complicated cases (38.3%) than females (20.8%). Complicated cases tend to present after 48 hours. The commonest postoperative complication was wound infection (5.5%). The mean length of hospital stay was 2.47 days, with no death was reported in the study period.

**Conclusion:** Peak appendicitis in summer is linked to rainfall and sunlight hours, affecting the 11-20 age groups.

## Introduction

Appendicitis, first described in 1886 by the pathologist Reginald Fitz(1), is defined as inflammation of the vermiform appendix that may lead to an abscess, ileus, peritonitis, or death if untreated(2). It is the most common surgical emergency in children and young adults with abdominal pain(3). The lifetime risk of developing acute appendicitis is 8.6% in males and 6.7% in females(4). The diagnosis of acute appendicitis is mainly a clinical one supported by laboratory investigation and imaging with most patients presenting with a typical history and signs, but a few may present in a typical manner(5). More than half of patients with acute appendicitis initially suffer mid-abdominal discomfort, which ultimately becomes more confined to the right lower abdomen area and is associated with a low-grade fever, nausea, vomiting, and anorexia(6). Cases may be simple (non-perforated) or complex (gangrenous, perforated, and pelvic/abdominal abscess)(7). Laparoscopic and open appendectomy are both safe and effective procedures for the treatment of acute appendicitis(8). Antibiotics are the first-line management for appendiceal masses or abscesses and have recently attracted a lot of attention in the treatment of non-perforated appendicitis(9).

The etiology of the condition remains uncertain; however, recently there is growing evidence that the occurrence of acute appendicitis displays seasonal disparity; more cases of appendicitis were noted during the summer, with monthly incidence correlated positively with the temperature(10). Surgeons continue to face an unpredictable situation regarding how many cases of appendicitis patients will visit emergency clinics each day, with some days bringing several cases and others bringing relatively few(11).

Ferris et al.’s (2017) study on the global incidence of appendicitis showed an incidence of 100 per 100,000 person-years in Northern America, and the incidence has been increasing in newly industrialized countries in Asia, Southern America, and Africa since 2000. Additionally, the paucity of population-based studies on the incidence of appendicitis in developing countries highlights a major gap in the literature(12).

Adu and Birhanu’s (2021) study showed 46.95% of the pooled prevalence of acute appendicitis in Ethiopia, with the Tigray region ranked first (51.98%), Addis Ababa (49.53%), and the Oromia region (47.75%)(13).

Ethiopia is located in a tropical region, and there are three seasons: (Summer): June, July, and August are the summer months. (Autumn): September, October, and November, sometimes known as the harvest season. (Winter): December, January, and February are the dry seasons with frost in the morning (14).

This study intends to increase understanding of the etiology of acute appendicitis by looking at seasonal variations and associated factors in cases of acute appendicitis. Such a study has not been carried out in Ethiopia.

## Methods

### Study Setting

The study was conducted in the surgical department of St. Paul’s Hospital Millennium Medical College (SPHMMC) in Addis Ababa, Ethiopia, a prestigious state tertiary care hospital.

### Study Period

The study was conducted from 01/09/2023 to 01/12/23 G.C for which a secondary data from patients’ charts whom were admitted from 01/01/2022 to 31/12/ 2022 G.C was utilized.

### Study Design

A cross-sectional study analyzing patient records with confirmed appendicitis and hospital discharge diagnosis of acute appendicitis and examined associations with weather variables, age, sex, and treatment outcome. Annual weather information was obtained from the Ethiopian National Meteorology Agency.

### Study Population

Patients admitted and received treatment for Acute appendicitis at SPHMMC in the study period.

#### Inclusion Criteria

Acute appendicitis patients treated and discharged at SPHMMC.

#### Exclusion Criteria

- Patients who underwent incidental appendectomy.
- Suspected case of Acute appendicitis that ended up having a different diagnosis
- Incomplete patient records

### Sample Size

The study analyzed 384 patients treated for acute appendicitis at SPHMMC from 01/01/2022 to 31/12/ 2022 G.C, for those meeting inclusion criteria. The sample size was calculated using single population proportion.

### Sampling procedures

Non-probability convenience sampling technique with quota assortment of the sample

### Measurement

#### Study Variables

##### Dependent Variable

- Acute Appendicitis

##### Independent Variables

- Seasons (average rainfalls/mm, average temperatures/degree Celsius, average sunlight/hours)
- Demography (age, sex)
- Intra-operative finding
- Postoperative complications

### Instruments

A questionnaire was used to gather patient information, including sociodemographic, months, symptoms, physical findings, complications, and disease-related factors, with weather variables from the Ethiopian National Meteorology Agency.

### Data Collection and Procedures

Trained data collectors surveyed patient charts and surgery registers between 01/09/2023 to 01/12/23 G.C, focusing on appendicitis diagnosis, surgical intervention, and treatment results.

### Operational Definitions

- Seasonal Variations: a systematic periodic occurrence of events for a year.
- Typical Appendicitis: related medical history was defined as abdominal pain which progressed from the peri-umbilical area to the RLQ and which was followed by either anorexia, nausea, or vomiting.
- Atypical Appendicitis: related medical history was defined as sudden, non-progressive abdominal pain, vague or absent pain localization, or predominant symptoms of diarrhea or vomiting.
- Simple appendicitis: cases of acute appendicitis that are self-limiting, non-perforated, or respond to antibiotics alone.
- Complicated appendicitis: perforated and gangrenous appendicitis or appendicitis with abscess or phlegmon formation.
- Leukocytosis: laboratory finding of WBC >10,000/mm3.
- Treatment outcome: The condition of the patient at discharge either improved and had no operative complication (favorable outcome) or improved but developed one or more complications or death (unfavorable outcome).
- Surgical Site infection: infection in the tissues of the incision and operation area.

### Data Quality Control and Management

Data quality was ensured through patient cards, surgical registration books, cross-matching, training sessions, questionnaire checks, and supervisors’ completion of data and code cleaning for improved reliability.

### Data Analysis

The study used SPSS 26.0 to analyze data, using the Shapiro-Wilks test for normality tests of continuous variables, the student’s test to compare mean ages, chi-square to compare nominal data, and Mann-Whitney and Kruskal-Wallis to compare simple and complicated appendicitis among groups and seasons. Pearson’s correlation coefficient was used to determine the correlation between monthly prevalence and weather variables, and Poisson regression was used to predict the degree of association between monthly prevalence and weather variables. Predictors of treatment outcomes for acute appendicitis analyzed by binary logistic regression. A p-value level of < 0.05 for significance.

### Ethical Consideration

The IRB of SPHMMC approved the study; patient data was safeguarded and confidentiality was maintained throughout the progress of the data collection process and patients’ charts were only utilized to this specific study.

## Results

### Socio-demographic magnitude and prevalence

The study analyzed 384 acute appendicitis admissions from 01/01/2022 to 31/12/2022 G.C, in the surgical department of SPHMMC, Addis Ababa, Ethiopia. Out of a total of 384 patients, 247 (64.3%) were males and 137 (35.7%) were females. The male-to-female ratio was almost 1.8:1. There were 7 patients admitted in pregnancy, 1^st^ trimester 1(0.3%) and 2^nd^ trimester 6 (1. 6%).The mean age was 20.8 years (SD = 13), the range was 2–83 years, and the median age was 18 years. A one-sample t-test was run to determine whether the mean age was different from a previous similar study of 20.5±9.7 SD (Ayele, 2021), which found that there was no statistically significant difference between means (t (383) =.433, p >.665), mean difference =0.297, 95% CL -1.05-1.65). With regards to catchment area, the study found that the majority of cases were from Burayu 115 (29.9%), Addis Ketema 107 (27.9%), and Gulele 50 (13.0%), with similar complicated cases from Burayu 79 (20.6%), Addis Ketema 52 (13.5%), and Gulele 22 (5.7%). The majority of the patients belonged to the second 128 (33.3%) and third decades 102 (26.5%) of life. Using the Chi square test, it was found that there was a significant association between patients’ age groups and sex in the occurrence of acute appendicitis (Chi-square x² = 11.910, df = 5, =*p*.036). The prevalence of acute appendicitis was higher among males in the age group 10–19 years 85(22.1%); similarly, among females, the prevalence was higher in the age group 10–19 years 43(11.2%). On the other hand, the prevalence of acute appendicitis has decreased with increasing age in both males and females.

**Table 1:**
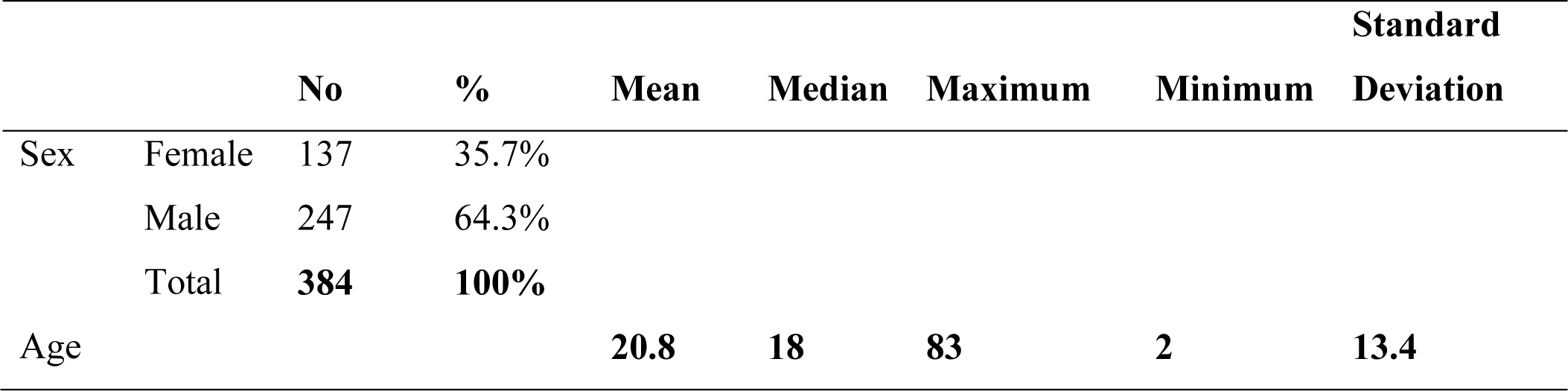
Sex and age distribution of acute appendicitis admissions among the study population during the study period.

|  |  | No | % | Mean | Median | Maximum | Minimum | Standard Deviation |
| --- | --- | --- | --- | --- | --- | --- | --- | --- |
| Sex | Female | 137 | 35.7% |  |  |  |  |  |
|  | Male | 247 | 64.3% |  |  |  |  |  |
|  | Total | **384** | **100%** |  |  |  |  |  |
| Age |  |  |  | **20.8** | **18** | **83** | **2** | **13.4** |

**Table 2:** Showing the prevalence of acute appendicitis by catchment area among the study population.

| Catchment area | Frequency | % |
| --- | --- | --- |
| Burayu | 115 | 29.9 |
| Addis ketema | 107 | 27.9 |
| Gulele | 50 | 13.0 |
| Kolfe | 46 | 12.0 |
| Arada | 12 | 3.1 |
| Holeta | 8 | 2.1 |
| Adaberga | 6 | 1.6 |
| Yeka | 6 | 1.6 |
| Sebeta | 5 | 1.3 |
| Lideta | 3 | 0.8 |
| Metarobi | 3 | 0.8 |
| Asko | 3 | 0.8 |
| Bole | 3 | 0.8 |
| Enchini | 3 | 0.8 |
| Munagusha | 2 | 0.5 |
| Asomeda | 2 | 0.5 |
| Sululta | 2 | 0.5 |
| Ambo | 1 | 0.3 |
| Debre Birhan | 1 | 0.3 |
| Guje | 1 | 0.3 |
| Gurshola | 1 | 0.3 |
| Jibat | 1 | 0.3 |
| Kaliti | 1 | 0.3 |
| Kirkos | 1 | 0.3 |
| Lafito | 1 | 0.3 |
| **Total** | **384** | **100.0** |

**Figure 1:**
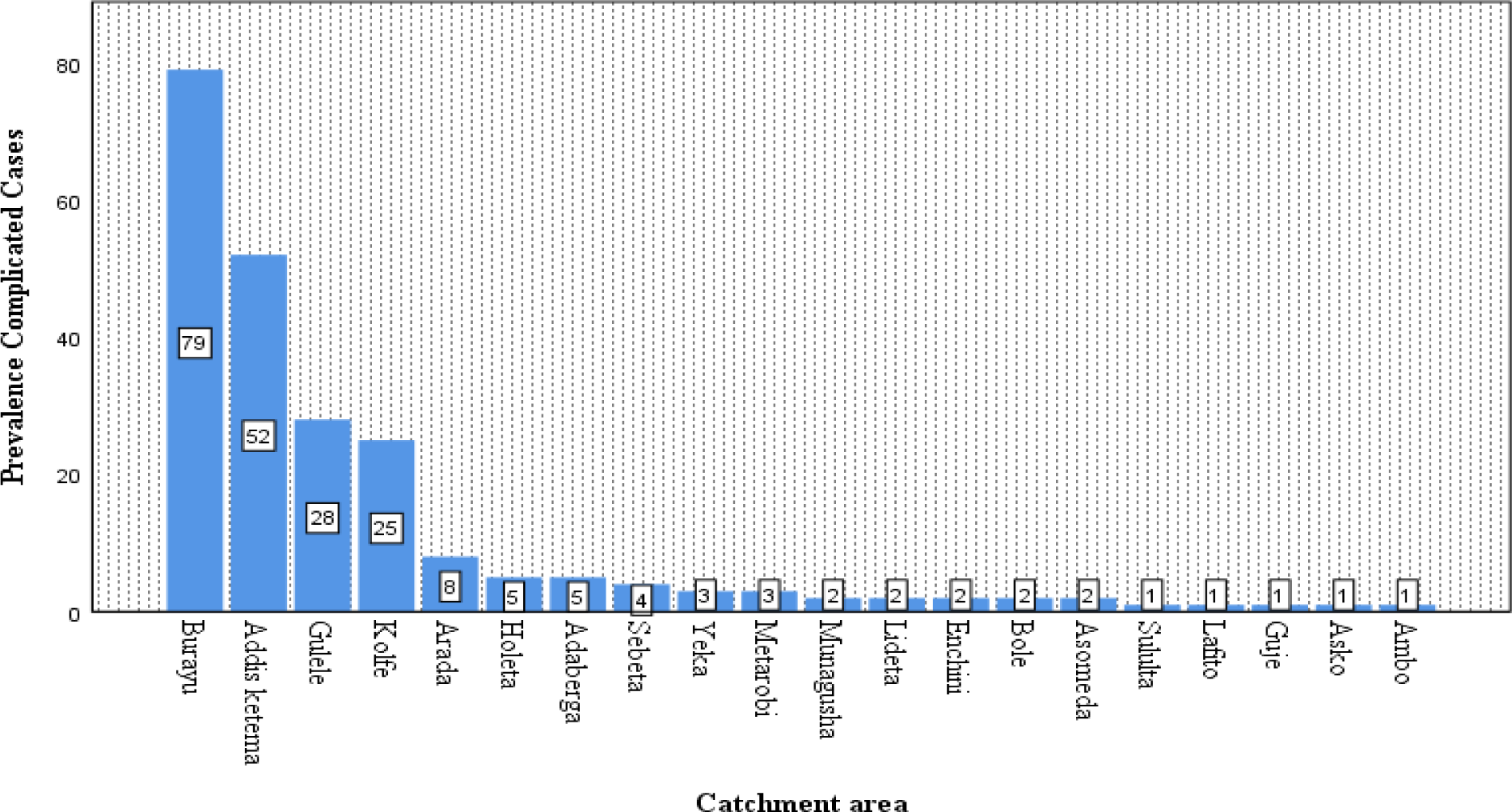
Showing Prevalence of complicated appendicitis by catchment area among study population.

**Table 3:**
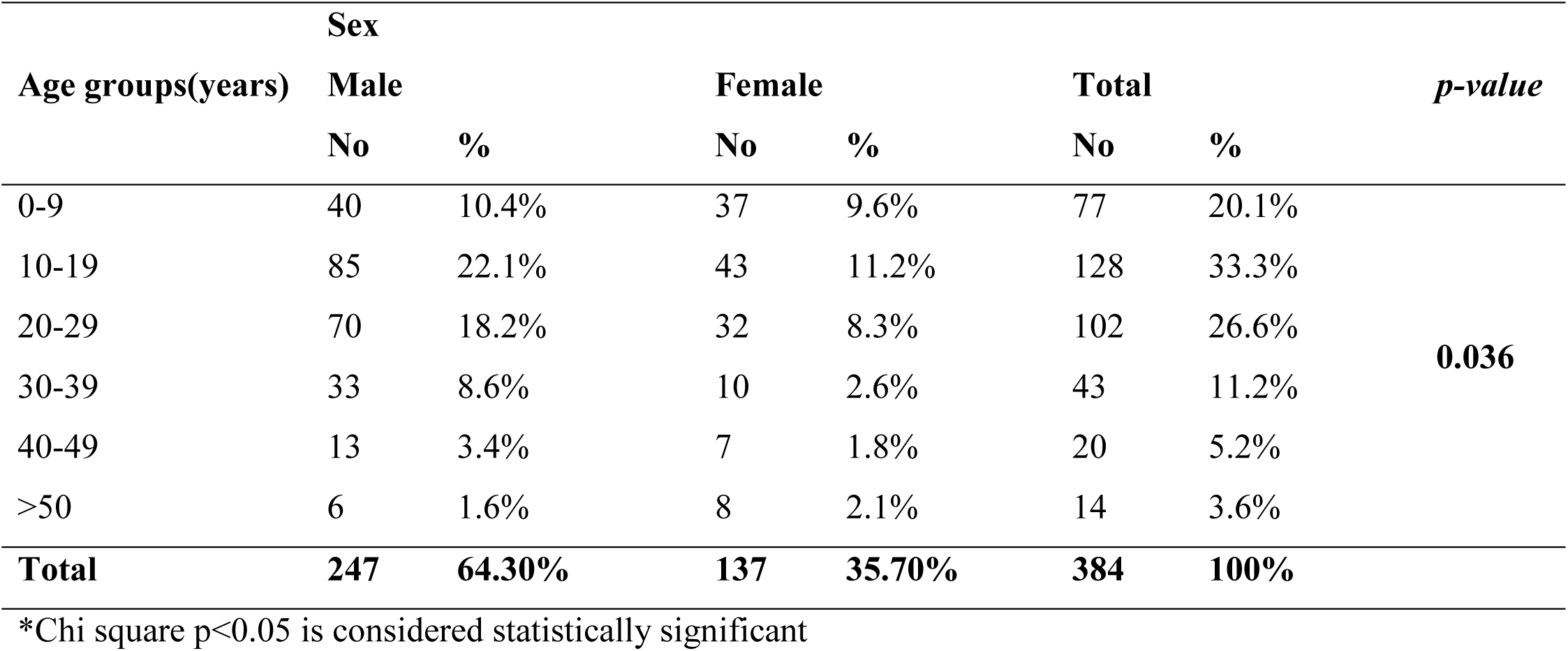
Association between prevalence of acute appendicitis, age group, and sex among the study population.

| Age groups(years) | Sex |  |  |  |  |  | *p-value* |
| --- | --- | --- | --- | --- | --- | --- | --- |
|  | Male |  | Female |  | Total |  |  |
|  | No | % | No | % | No | % |  |
| 0-9 | 40 | 10.4% | 37 | 9.6% | 77 | 20.1% | **0.036** |
| 10-19 | 85 | 22.1% | 43 | 11.2% | 128 | 33.3% |  |
| 20-29 | 70 | 18.2% | 32 | 8.3% | 102 | 26.6% |  |
| 30-39 | 33 | 8.6% | 10 | 2.6% | 43 | 11.2% |  |
| 40-49 | 13 | 3.4% | 7 | 1.8% | 20 | 5.2% |  |
| >50 | 6 | 1.6% | 8 | 2.1% | 14 | 3.6% |  |
| **Total** | **247** | **64.30%** | **137** | **35.70%** | **384** | **100%** |  |

\*Chi square $p < 0.05$ is considered statistically significant

### Pattern of clinical features

All patients had an initial attack of abdominal pain, which either became localized to right lower quadrant or diffuse abdominal pain, followed by anorexia in 378 (98.4%), nausea and vomiting in 336 (87.7%), and fever in 231 (60.2%). The important physical findings were right lower quadrant tenderness 340 (88.5%), diffuse abdominal pain in 32 (8.3%), and RLQ mass 10 (2.6%). The key associated symptoms at presentation were diarrhea in 24 (6.3%), URTI in 18 (4.7%), and dysuria in 12 (3.1%). At diagnosis, the mean pre-operative WBC count was 13500 cells/mm3 (SD = 4.9) and ranged from 3000–3200 cells/mm3. The mean granulocyte count was 81% (SD = 9.2) and ranged from 45%–96%.

**Table 4:**
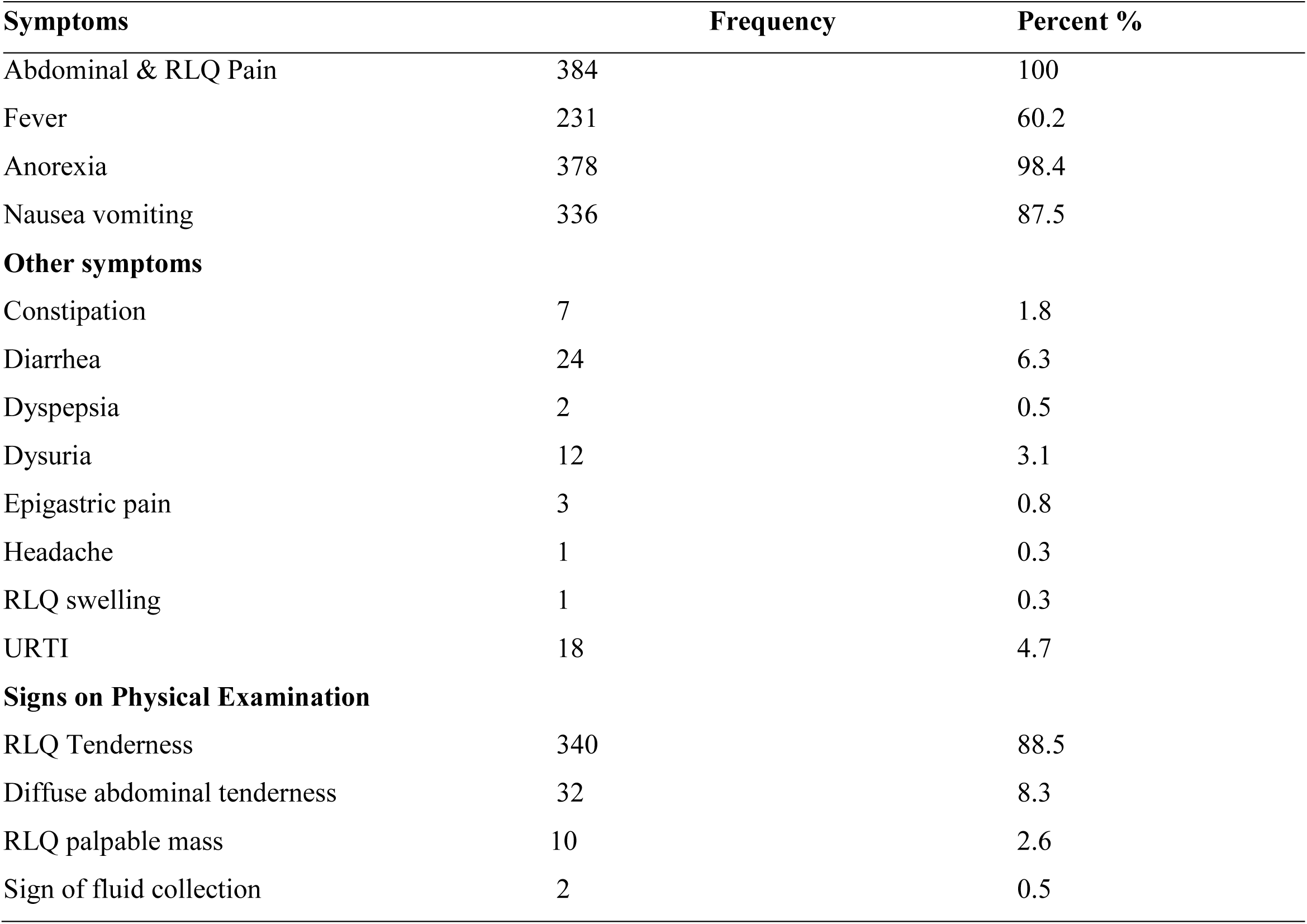
Distribution of clinical symptoms and signs of acute appendicitis cases among study population from 01/01/2022 to 31/12/2022 G.C (N=384)

| Symptoms | Frequency | Percent % |
| --- | --- | --- |
| Abdominal & RLQ Pain | 384 | 100 |
| Fever | 231 | 60.2 |
| Anorexia | 378 | 98.4 |
| Nausea vomiting | 336 | 87.5 |
| **Other symptoms** |  |  |
| Constipation | 7 | 1.8 |
| Diarrhea | 24 | 6.3 |
| Dyspepsia | 2 | 0.5 |
| Dysuria | 12 | 3.1 |
| Epigastric pain | 3 | 0.8 |
| Headache | 1 | 0.3 |
| RLQ swelling | 1 | 0.3 |
| URTI | 18 | 4.7 |
| **Signs on Physical Examination** |  |  |
| RLQ Tenderness | 340 | 88.5 |
| Diffuse abdominal tenderness | 32 | 8.3 |
| RLQ palpable mass | 10 | 2.6 |
| Sign of fluid collection | 2 | 0.5 |

### Pattern of intraoperative findings among the study population

The majority of cases had classical appendectomy 311 (81%), while laparotomy was done in 73 (19%). Complicated cases accounted for the majority of the patients, 227 (50.9%), which include intra-abdominal abscess 66 (17.2%), phlegmon 63(16.4%), peri-appendiceal abscess 43 (11.2%), perorated 41 cases (10.7%), gangrenous 9 (2.3%), and appendiceal mass 5 (1. 3%).The uncomplicated cases accounted for a lesser number, 157 (40.9%).

**Figure 2:**
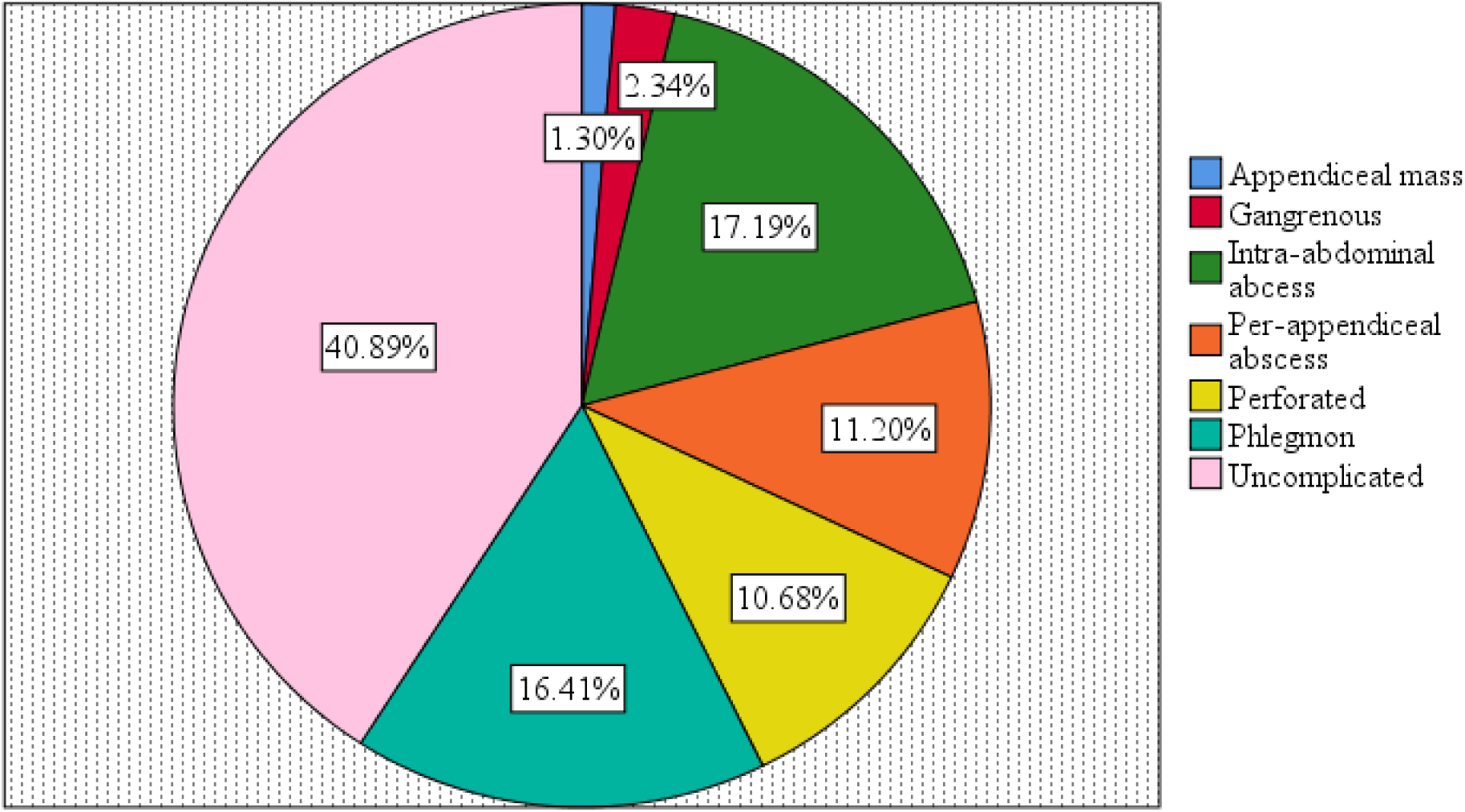
Displays the pattern of intra-operative findings in cases of acute appendicitis.

### Effects of Season and weather variables on prevalence of Acute Appendicitis

In the study, higher admissions of patients with acute appendicitis were in rainy months, with a gradual, minimal increment starting in the autumn months of March 8.6% (33), April 8.1% (31), and May 7.3% (28) and peaking in the summer months (32.8% of all, P < 0.05) of June 9.9% (38), July 12.5% (48), and August 10.4% (40). Then it starts with a minimal decline from the spring months of September 6.3% (24), October 6.3% (24), and November 8.1% (31) and the winter period of December 6.3% (24), January 8.1% (31), and February 8.3% (32). The slightly lower prevalence was in September and October.

**Figure 3:**
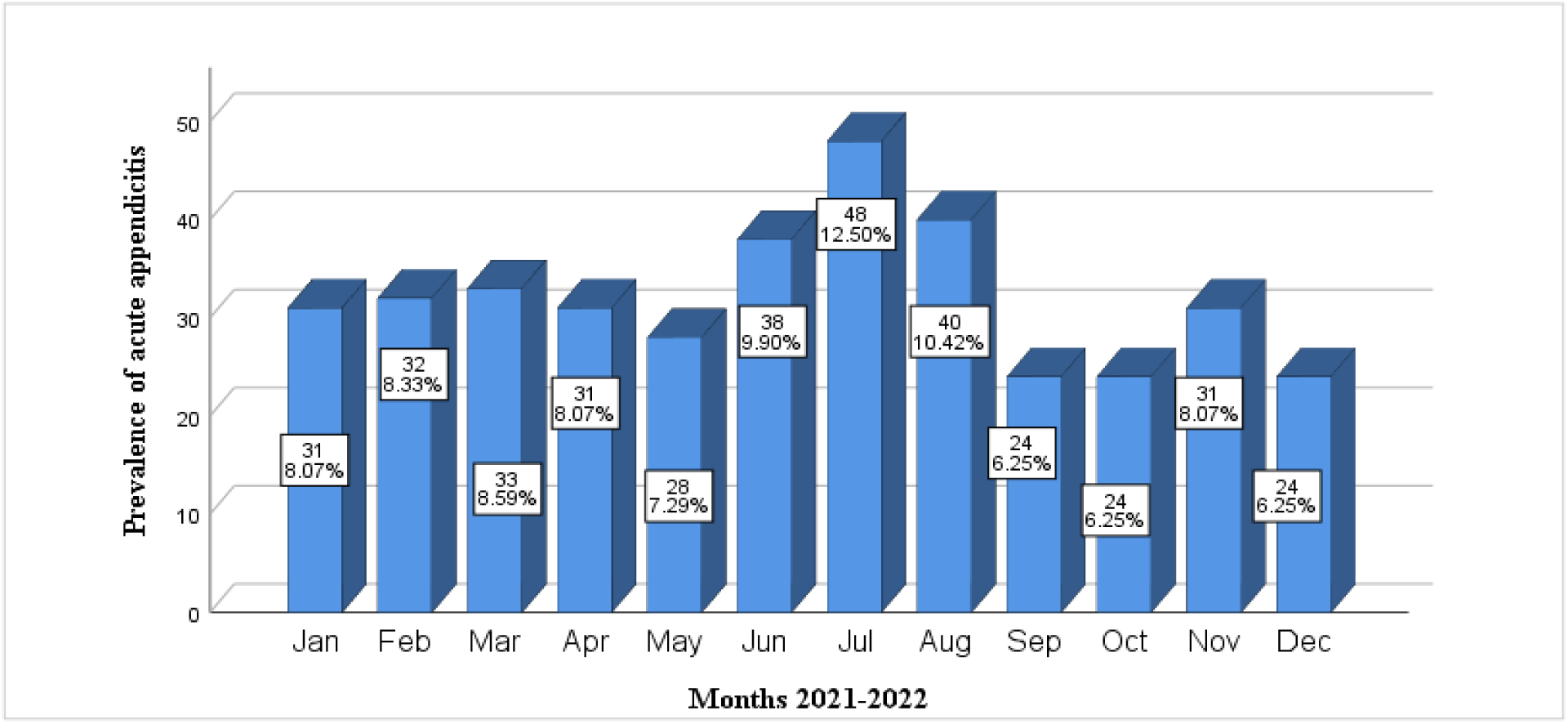
Displays cumulative prevalence of acute appendicitis admissions from 01/01/2022 to 31/12/2022 G.C resolved by months.

### Rainfall and Rainy days

There is a notable fluctuation in precipitation, with a 412 mm variance observed between the month of least rainfall in December (7mm) and that of highest downpour in August (419mm). The summer months (June 271mm, July 386mm, and August 491) are the period of highest precipitation (wet-2 season in Addis Ababa). The wet-1 season is the autumn months (March 72mm, April 147mm, May 171mm) in Addis Ababa. The summer heavy rainy months were found to coincide with the highest prevalence of acute appendicitis associated with the highest episodes, 126 (32.81%) overall cases. Likewise, the number of rainy days per month has a wide fluctuation with a range of 20 days; the rainiest number of days was observed in the summer months in June (17 days), July, and August (22 days), which was found to coincide with the highest episode 126 (32.8%) overall cases of acute appendicitis. The lowest rainy days occurred in the winter months: December (1 day), January (3 days), and February (4 days), which registered the lowest episodes of acute appendicitis in 87 (22.7%) overall cases. When Pearson product correlation coefficient testing was performed, average rainfall/mm, rainy days, and prevalence of acute appendicitis were found to be moderately positively correlated and statistically significant (r =.637, *p* <.05), suggesting that increased rainfall and rainy days may raise the prevalence of acute appendicitis.

**Figure 4.**
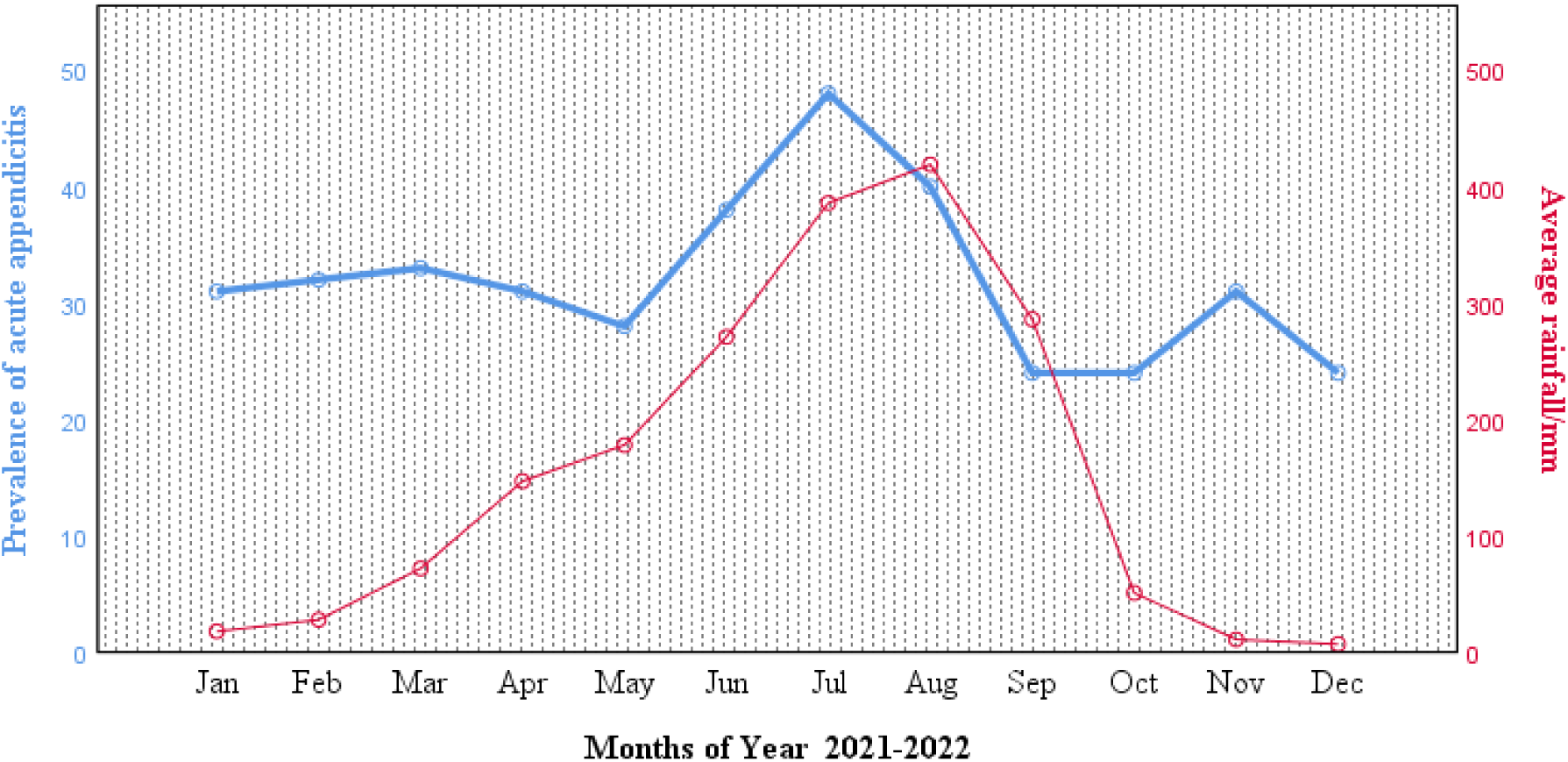
: Displays cumulative prevalence of acute appendicitis admissions from 01/01/2022 to 31/12/2022 G.C against the average monthly rainfall/mm for the same period and geographic area.

### Sunlight hours

As could be expected, with a range of 3.9 hours (maximum 9.8 hours and minimum 5.9 hours), the average sunlight hours were found to be lowest during the summer months August (5.9 hours), July (6.1 hours), June (7.8 hours), and highest during the winter (December (9.3 hours), January (9.3 hours), February (9.8 hours), and autumn months (March (9.6 hours), April (9.5 hours), and May (9.3 hours), respectively. The summer months with the lowest sunlight hours were found to correspond with higher rates of acute appendicitis: June 38 (9.9%), July 48 (12.5%), and August 40 (10.42%). The months with higher average sunlight hours recorded a lower rate of acute appendicitis. When Pearson product correlation coefficient testing was performed, average sunlight hours and the prevalence of acute appendicitis were found to be **moderately negatively correlated** and statistically significant (r = -0.618, *p* <0.05), suggesting that decreased sunlight hours may raise the prevalence of acute appendicitis.

**Figure 5:**
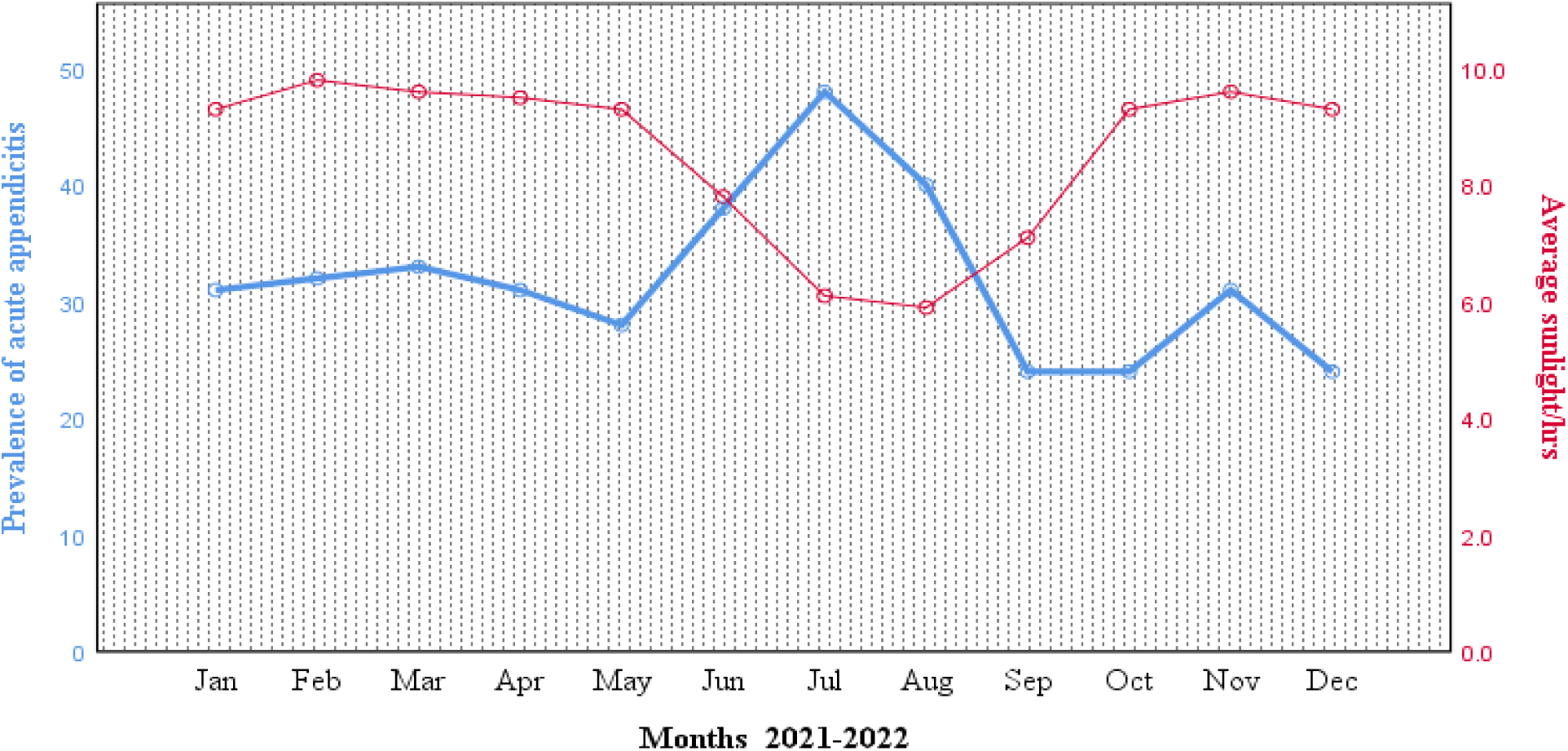
Displays cumulative prevalence of acute appendicitis admissions from 01/01/2022 to 31/12/2022 G.C against the average number of monthly sunlight hours for the same period and geographic area.

### Temperature

The temperature in Addis Ababa showed no much fluctuations. The variation in temperatures throughout the year is 2.8 °C. On average, the month of April experiences the highest temperature, with an average value of 17.2 °C. The lowest average temperatures in the year occur in December, when it is around 14.4 °C. Lower temperatures occur in summer, spring, and partly winter, while the hotter months are in autumn. A 2 °C drop in temperature observed from autumn to summer initially showed an increase in episodes of acute appendicitis in summer temperature; however, further decreases in temperature through September (14.8 °C), October (14.6 °C), November (14.7 °C), and December (14.4 °C) registered lower rates of acute appendicitis. When Pearson product correlation coefficient testing was performed, average monthly temperatures and the prevalence of acute appendicitis found negligible negative correlation (r = -0.037, *p* >0.05).

**Figure 6:**
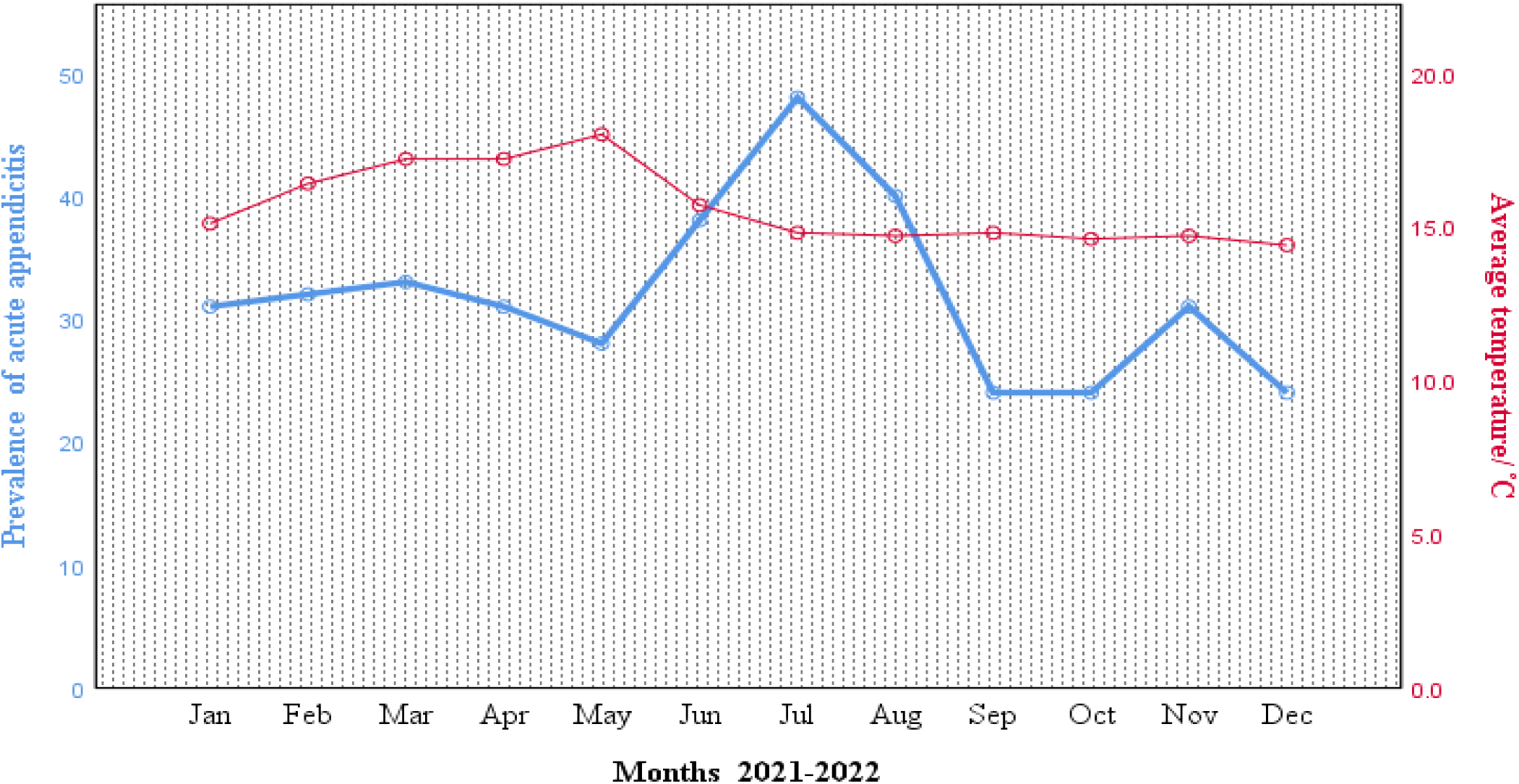
Displays cumulative prevalence of acute appendicitis admissions from 01/01/2022 to 31/12/2022 G.C against the average number of monthly temperature/°C for the same period and geographic area.

### Humidity

The month with the highest relative humidity is August (87%). The month with the lowest relative humidity is February (44%). Relative humidity is very high in the summer months of June (76%), July (86%), and August (87%), which corresponds with a higher occurrence of acute appendicitis in the summer 126 (32.8%). The relative humidity is lowest in the winter months: December (52%), January (525%), and February (44%), which recorded fewer cases of acute appendicitis in 87 (22.7%) overall cases. When Pearson product correlation coefficient testing was performed, average monthly humidity and the prevalence of acute appendicitis found a negligible positive correlation (r =0.57, *p* >0.05).

**Figure 7:**
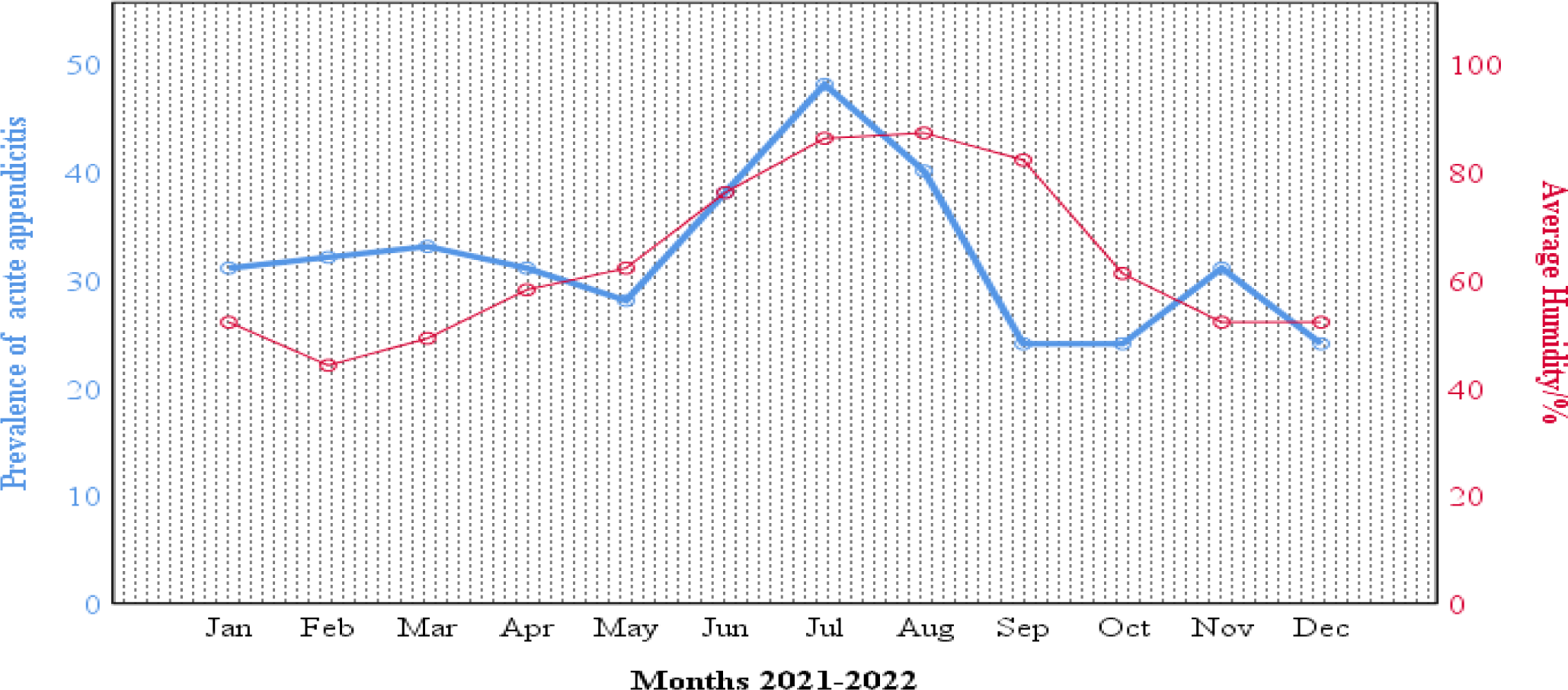
Displays cumulative incidence of acute appendicitis admissions from 01/01/2022 to 31/12/2022 G.C against the average relative humidity/% for the same period and geographic area.

**Table 5:**
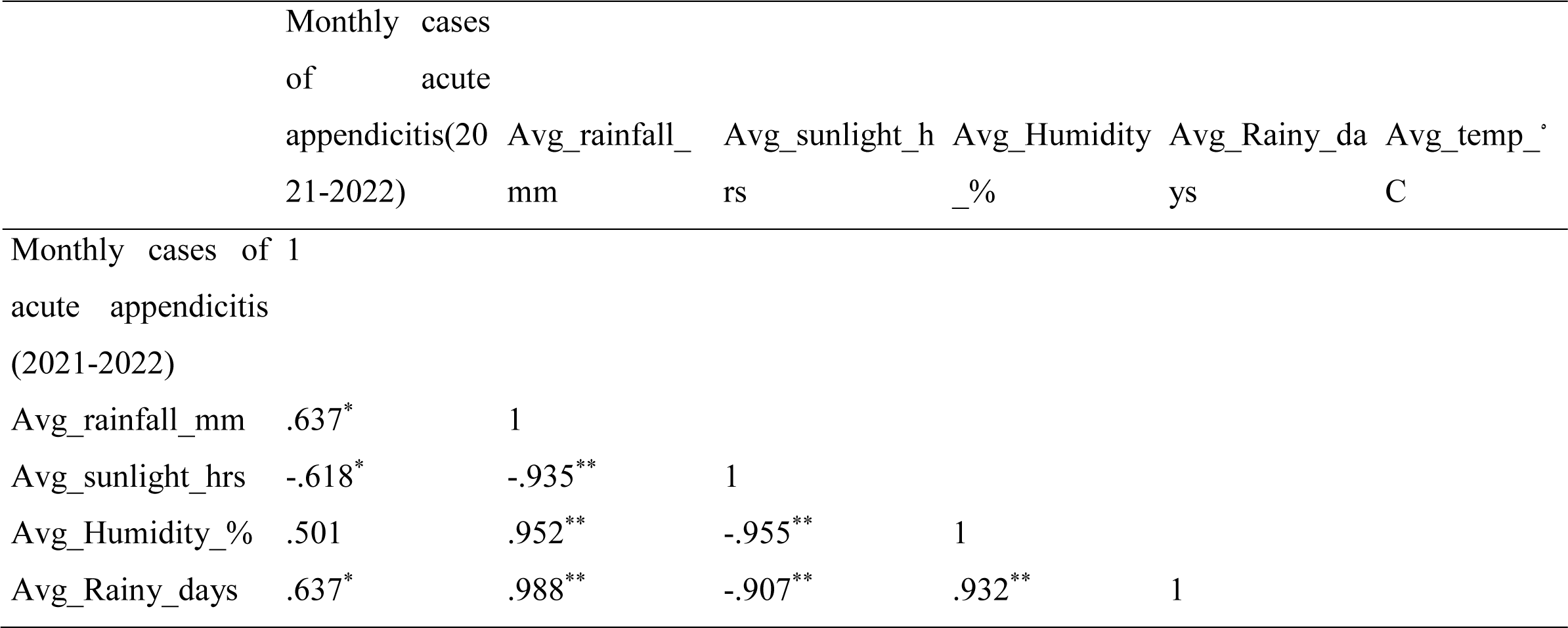
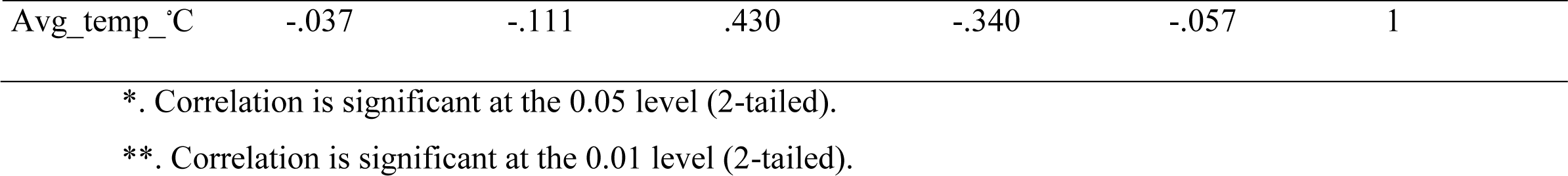
displays Correlation analysis matrix of the monthly prevalence of acute appendicitis and average recorded weather variables for the same period and geographic area.

### Poisson regression analysis between monthly prevalence of acute appendicitis and average recorded monthly weather variables

The study used **Poisson regression analysis** to determine if weather variables (average rainfalls, sunlight hours, rainy days, sun light hours, relative humidity, and temperatures) could predict acute appendicitis incidence. Results showed a significant improvement in fit over a null model, but no weather variable significantly predicted the monthly appendicitis prevalence.

**Table 6:** Shows Omnibus test for the full model significance.

| **Omnibus Test<sup>a</sup>** |  |  |
| --- | --- | --- |
| Likelihood Ratio Chi-Square | df | Sig. |
| **11.455** | **5** | **.043** |

Dependent Variable: Montly\_cases\_2021\_2022
Model: (Intercept), avg\_rainfall\_mm, avg\_sunlight\_hrs, avg\_Rainy\_days, avg\_Humidity\_%, avg\_temp\_°C
a. Compares the fitted model against the intercept-only model.

**Table 7:**
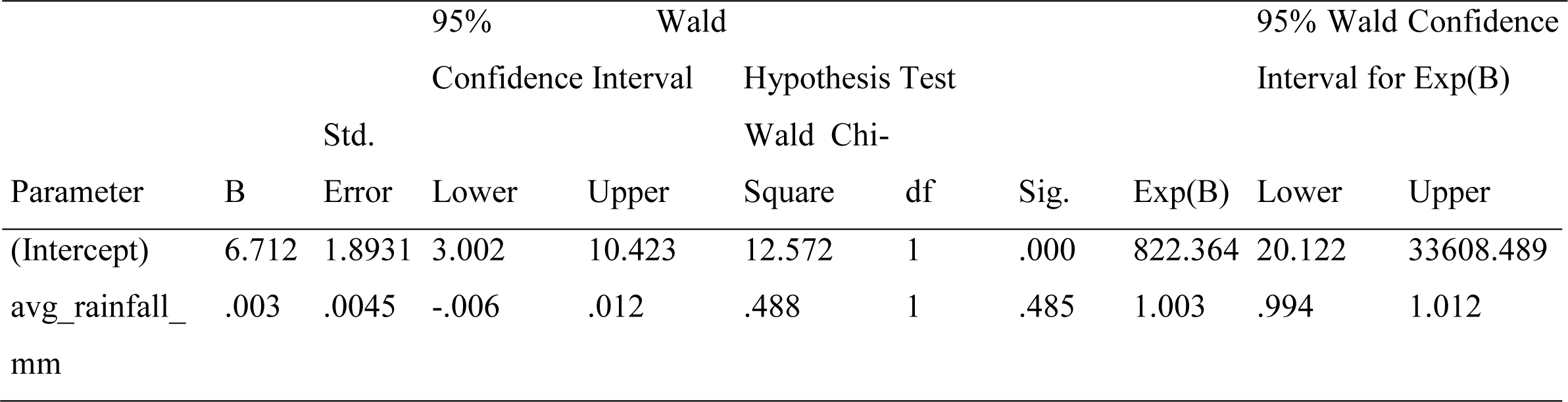
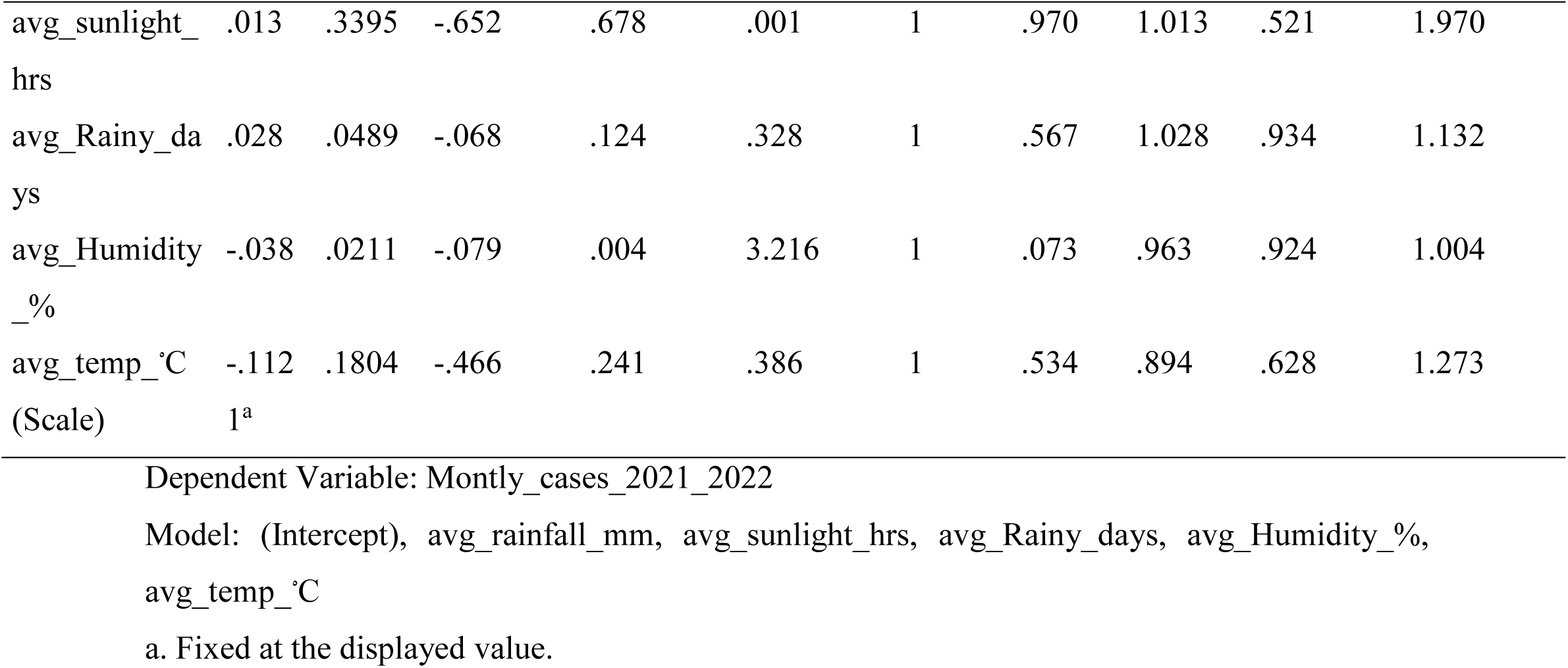
Poisson Regression analysis matrix showing parameter estimates.

### Seasonal prevalence of complicated and Simple cases of acute appendicitis

Through our study, we also aimed to investigate which season of the year complicated cases present more. Out of 227 cumulatively complicated cases, the study revealed that summer had the highest number of complicated cases, 79 (34.8.4%), followed by spring 51 (22.5%), autumn 50 (22.0%), and winter 47 (20.7%), with a low complication rate. The **Kruskal-Wallis test** performed showed no statistically significant difference in the distribution of complicated cases across the seasons (T = 3.00, df = 3, *p* = 0.392).

**Figure 8:**
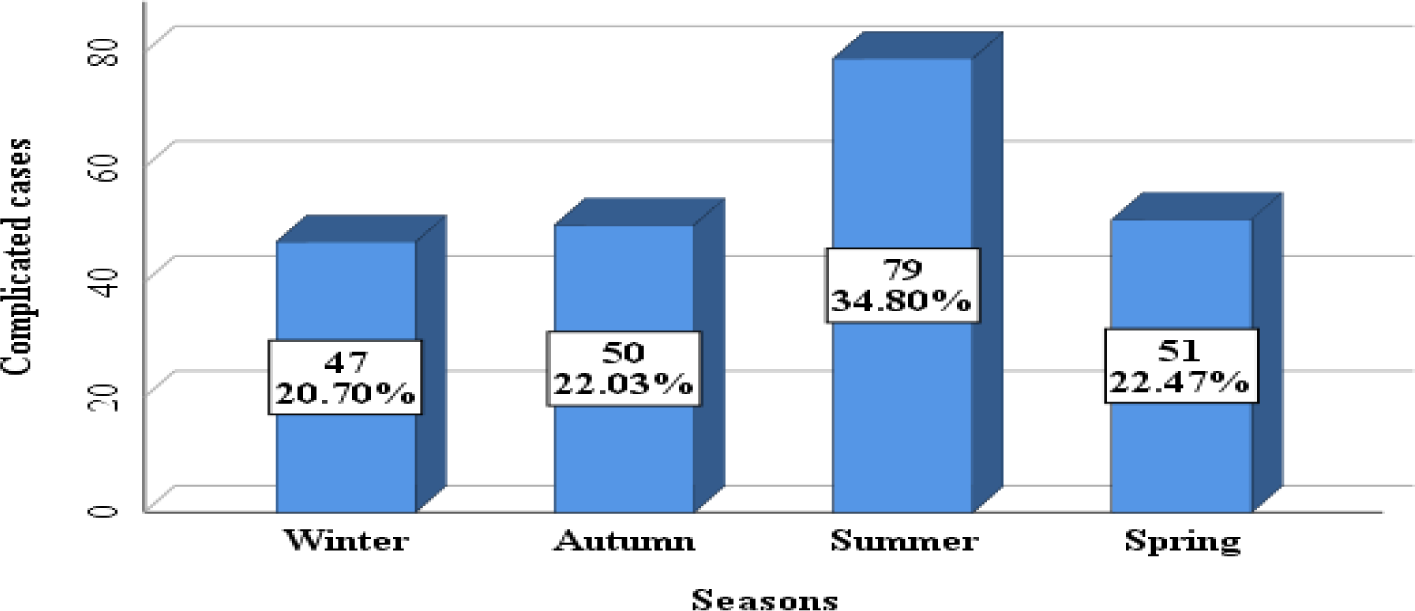
Seasonal prevalence of complicated and Simple acute appendicitis.

### Association of determinant variables and surgical treatment outcome of acute appendicitis Preoperative laboratory WBC parameters

At diagnosis, the mean pre-operative WBC count was 13500 cells/mm3 (SD = 4.9) and ranged from 3000–3200 cells/mm3. The mean granulocyte count was 81% (SD = 9.2) and ranged from 45%–96%. **A Mann-Whitney U test** was performed to evaluate the difference in preoperative laboratory WBC between complicated and simple cases. The test found that preoperative WBC values were significantly higher in complicated cases (median = 14.00, n = 227) compared to simple cases (median = 11.90, n = 157), U = 114009.5, Z = 3.565, *with an effect size of r = 0.18, p* = 0.00).

**Table 8:** Descriptive statistics of the preoperative laboratory parameters.

|  | Mean | Maximum | Minimum | Standard Deviation |
| --- | --- | --- | --- | --- |
| Preop_Lab_WBC_1000_cells/mm<sup>3</sup> | 13.5 | 32.0 | 3.0 | 4.9 |
| granulocyte% | 81.0 | 96.0 | 45.0 | 9.2 |

### Duration of illness compared with intraoperative findings among the study population

Majority of our patients presented in 12-24 hours 147(38.3%), followed by 48 hours 122(31.8%) with least patients presenting in less than 12hour 43(11.2%). Common duration of presentation among complicated cases was after 48hours 106 (27.6%) with least complication rate in those presenting less than 12 hours 9 (2.3%). Using the **Chi square test**, it was found that there was a significant association between duration of illness and complication rates (Chi-square x² = 79.498, df = 3, *p* > *0.00)*.

**Table 9:**
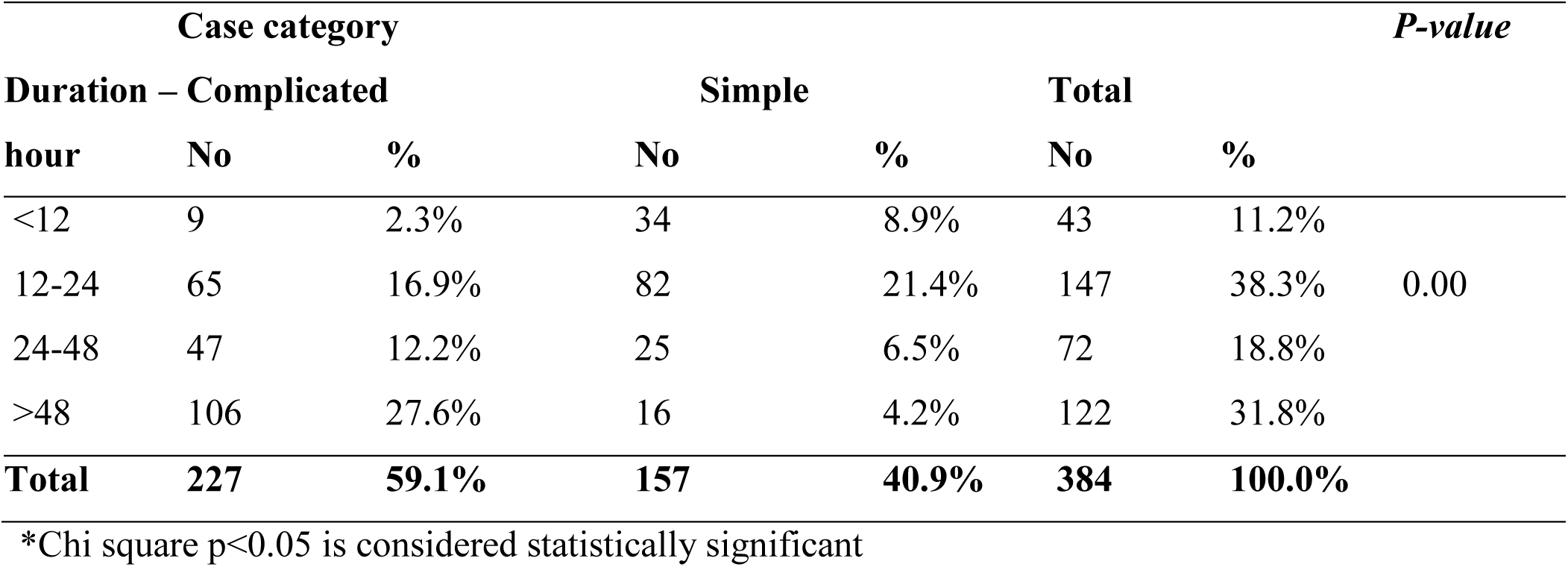
Displays Association between duration of illness and complication rates among the study population.

### Age and sex distribution of uncomplicated and simple acute appendicitis among the study population

Complicated cases made up 59.1% (227), while simple cases made up 40.9% (157). The 10–19 age group had 82 (21.3%) more complications. Simple cases were also observed to be slightly higher (49, 12.8%) in the 20–29 age group. Males were responsible for a somewhat higher proportion of complicated cases (147, 38.3%) than females (80, 20.8%). Males likewise had a higher proportion of simple cases (100%) than females (57, 14.8%). The study found no statistically significant differences in complication rates by age (**Chi-square** x² = 6.359, df = 5, *p*=0.273) or sex (Chi-square x² = 0.046, df = 1, *p=* 0.831).

**Table 10:**
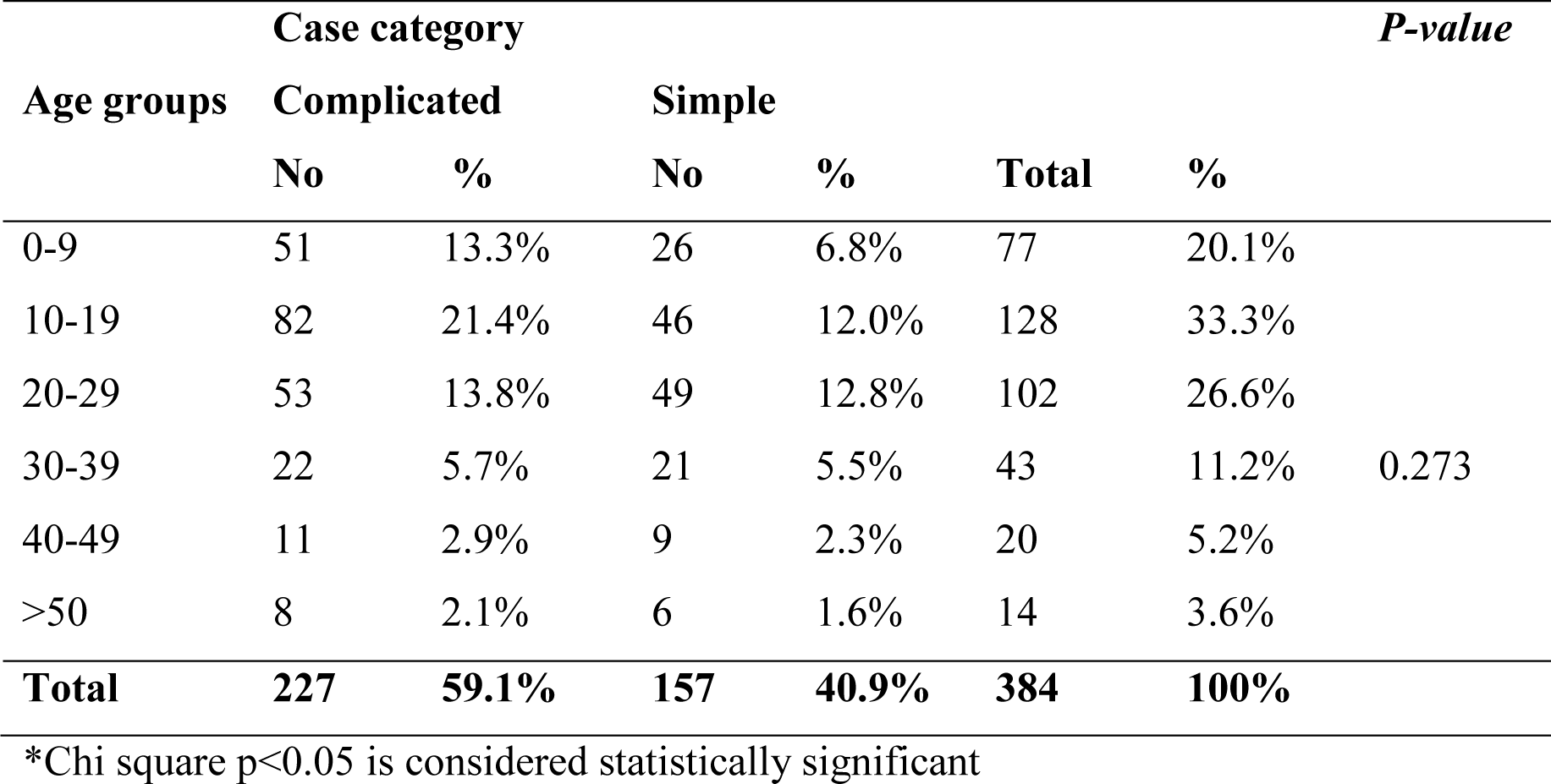
Shows Age groups distribution of complicated and simple acute appendicitis.

**Table 11:**
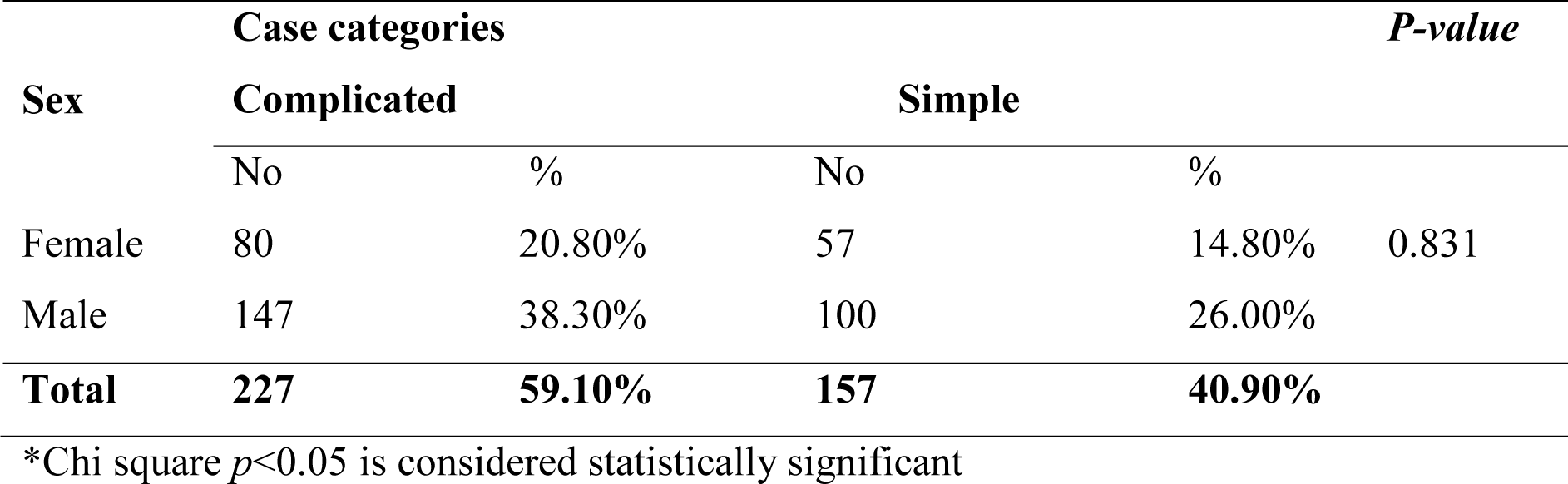
Shows sex distribution of complicated and Simple cases of acute appendicitis.

|  | Case categories |  |  |  | *P-value* |
| --- | --- | --- | --- | --- | --- |
| Sex | Complicated |  | Simple |  |  |
|  | No | % | No | % |  |
| Female | 80 | 20.80% | 57 | 14.80% | 0.831 |
| Male | 147 | 38.30% | 100 | 26.00% |  |
| Total | 227 | 59.10% | 157 | 40.90% |  |

\*Chi square $p < 0.05$ is considered statistically significant

### Length of hospital stays, post-operative complication rate, and predictors of treatment outcomes for acute appendicitis among the study population

In the study, there were no hospital deaths registered due to acute appendicitis, and the overall mean length of hospital stay was 2.47±(SD=2.48) days (range: 1-27 days) with complicated cases 3 ±(SD=3.0) days mean length of hospital stay and simple cases with mean length of stay 1 ±(SD=1.0) day. **A Mann-Whitney U test** was performed to evaluate the difference in hospital length of stay between complicated and simple cases. The test found that hospital length of stay was significantly higher in complicated cases (median = 3.00, n = 227) compared to simple cases (median = 1.00, n = 157), U = 5669.00, Z = 12.462, p =.00, with an effect size of r =0.5. Most of the patients (349) (90.9%) had no postoperative complications, and 35 (9.1%) had postoperative complications. The commonest postoperative complication was wound infection (5.5%), followed by sepsis (2.6%). Wound infections were common in both the age groups 0–9, 5 (1.3%) and 10–19, 5 (1.3%). 8 (2.1%) cases were re-explored due to intra-abdominal sepsis, and 1 case (0.35%) had ICU admission due to sepsis.

**Table 12:** Displays frequency of post-operative complications cases of acute appendicitis among the study population.

| Postoperative complication | No | % |
| --- | --- | --- |
| Respiratory | 4 | 1.0 |
| Sepsis | 10 | 2.6 |
| Wound infection | 21 | 5.5 |
| **Total** | **35** | **9.1** |

The determinant variables that were found to predict the favorable or unfavorable treatment outcome of cases of acute appendicitis were found to be statistically associated in binary logistic analysis. Hospital length of stay was associated with a predicted good treatment outcome (OR = 468 at 95% CI 0.363–0.603, p-value = 0.00), with a 0.177% deduction in hospital length of stay. Duration of presentation to the hospital after illness for less than 12 hours was associated with a favorable treatment outcome (OR 3.870 at 95% CI 1.363–10.988, P-value = 0.011), with 3.87% having a favorable outcome compared to an unfavorable treatment outcome. Low values of white cell counts were associated with having simple appendicitis (OR 0.446 at 95% CI 0.446-0.938, P-value = 0.022), with a 0.647% chance of simple appendicitis compared to complicated appendicitis. Sex and age were not significant predictors of treatment outcome, respectively.

**Table 13:**
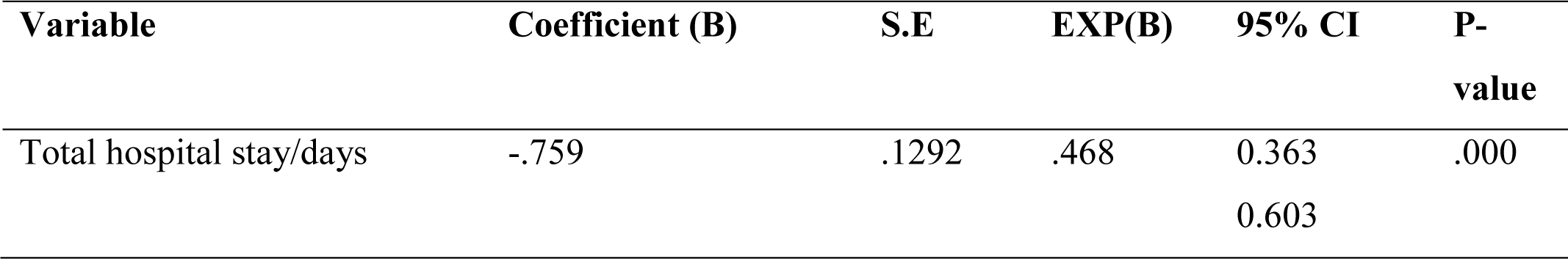
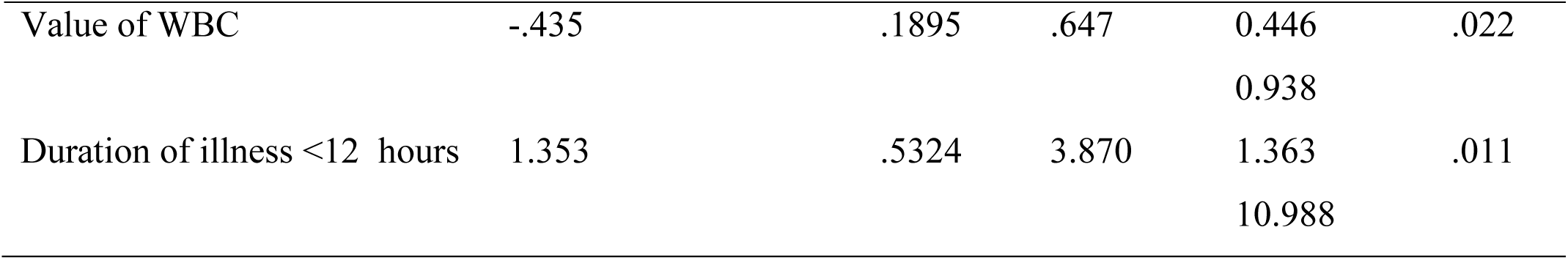
Shows predictors of treatment outcome of acute appendicitis among study population.

## Discussion

The demographic facts from 384 admission events supports previous studies, with male predominance (64.3% male, 35.7% female), and a male-to-female ratio of 1.8:1, and a mean age of 20.8. Which is similar to studies done at Wollo University in Dessie, Ethiopia by Ayele et al. (37)(38), in Pakistan(30),Turkey (39). The sample appears to be more comprehensively representative of patients with acute appendicitis. The study found a significant association between age and sex, with the majority of acute appendicitis cases occurring between 10 and 19 years old (33.3%), both males and females, and the least common at over (3.6%). Which could be explained by a higher incidence of acute appendicitis in this age group, which is consistent in Nigeria by Oguntola et al.(24),Yemen (26), Iraq (40), India (7), Global Burden of Disease Study 2019(21),Turkey (11). The study revealed no significant differences in complication rates based on age or gender. Acute appendicitis in pregnancy was more prevalent in 2nd trimester 6 (1.5%) than in 1st trimester 1 (0.3%), which is similar to the study done in Turkey (41). The study consistently reports a typical pattern of clinical symptoms, including abdominal pain, anorexia, vomiting, and fever, Jimma University, Ethiopia by Ayana et al. (37), USA(15).

Our study found that acute appendicitis is present throughout the year, but some particular months are associated with a higher occurrence. The study found a significant seasonal variation in cases of acute appendicitis, with a peak in Summer 126 (32.8%), compared to other seasons. The month with the highest number of cases (48) was July, coinciding with the year’s heavy rainfalls and bottom sunlight hours recorded. The prevalence of cute appendicitis was moderately positive and negatively correlated with rainfall and sunlight hours. Our findings align with previous studies conducted on this issue in Nigeria (24), Cameroon(23),UK (28), Pakistan(30), India(7), Bahrain(10), Nepal(27), Irag(29), Iran(32). other studies reported higher prevalence of acute appendicitis during the Winter and Autumn season in Yemen (26), winter and autumn in Turkey (39). Seasonal variation suggests potential heterogeneous extrinsic factors like rainfall, viral and bacterial infections, allergens, sun radiation, and dehydration could contribute to appendicitis etiogenesis (42), (30). The number of cases steadily increases from June due to the onset of the rainy season, intensifying in July and August. During this period, increased bacterial, viral, and parasitic infections, particularly in regions with high humidity, heavy rainfall, and poor sanitation, may contribute to an increased incidence of appendicitis. (43)(44), in Nigeria (24). Bagus et al.’s 2020 study in Indonesia revealed that 51.5% of pediatric appendicitis patients were linked to low hygiene (34).Some cases of lymphoid hyperplasia, an immunological response, may be attributed to allergic reactions to pollen from flowers like maize during the rainy season (45), (46). Dietary habits in summer might play a role. Dehydration, low-fiber diets, sugary foods, and high alcohol consumption can lead to constipation, infection, and appendicitis, especially during summer when summer is associated with the highest alcohol consumption (47). (48) (49). The study revealed common associative symptoms of appendicitis include diarrhea (6.3%), URTI (4.7%), and constipation (1.8%). However, further research is needed on trends in appendicitis incidence in populations consuming traditional diets with different fiber content. The study found that the role of temperature and relative humidity in seasonal variation was weaker, similar to the research conducted in Bahrain (10).

The postoperative hospital stay averaged 2.47 days, slightly shorter than previous studies’ 3.2 days done at our hospital (50). Postoperative hospital stays are significantly influenced by disease clinical stage (P = 0.000), with complicated cases requiring longer stays (3.36 days) compared to simple cases (1.2 days), a similar finding in India (7). Dagebo et al. at St. Luke Catholic Hospital in Woliso, Ethiopia, reported a 10.4-day duration for complications compared to a 6.6-day duration for simple cases (35). The study found that complicated cases (27.6%) typically present after 48 hours (p > 0.00), with illness duration significantly influencing intraoperative findings, similar to the study done in our hospital (50), in Woliso, Oromia, Ethipoia (35).

The study’s post-operative complication rate was 9.1%, comparable to the Wollo, Ethiopia (11.7%) (51), in South Africa urban population(52), kenya 12.3%(53). Postoperative complications include wound infection at 5.5%, intra-abdominal sepsis at 2.6%, and pneumonia at 1%, similar to South Africa’s 6.7% wound infection rate and 5% pneumonia rate (52).The Nigerian report showed a wound infection rate of 26.8% (54). The study found no fatalities, similar to those conducted at Kenyatta National Hospital, Kenya, and Wollo University, Ethiopia (51), (53). However, other studies, like in Nigeria, reported very low mortality (0.7%). South Africa: 0.3% (54), (52). The improvement is primarily attributed to enhancements in patient care, which includes enhanced preoperative protocols and postoperative care.

## Conclusions

In line with other studies, Acute Appendicitis is higher in males than females, with a male-to-female ratio of 1,8:1. The prevalence peaks in the teen-age-group in both sexes, due to a highly responsive lymphoreticular system. Late presentation and being young are associated with complications. The study found seasonal variation in appendicitis occurrence, with more admissions in the summer, with a peak in July and a low in September, October, and December. Factors contributing to this variation include infectious agents’ natural life cycles, diet, allergen exposure, and environmental factors.

## Recommendations

- The study suggests implementing extra preventive measures and raising community awareness about risk factors for acute appendicitis, particularly during the summer, to reduce the disease burden through the provision of safe water, good sanitation, and hygiene practices.
- Guidance at schools for children on a healthy diet, the significance of safe drinking water, and personal hygiene, as they are the most vulnerable to appendicitis.
- Adult need 2.5 liters of water per day. The daily diet should contain a sufficient amount of fiber especially during summer.
- The treating clinicians need to have a high index of suspicion of acute appendicitis for males and those in the 2nd and 3rd decades of life, especially during the summer.
- Further large-scale multicenter studies over several years are recommended to draw better conclusions on these observations.

## Limitations

- The study’s single-center survey population, primarily from Addis Ababa, may not accurately represent the broader context due to the majority of patients being from a specific location.

## Data Availability

All relevant data are within the manuscript and its Supporting Information files.

## Acknowledgement

Our sincere appreciation goes out to SPHMMC for giving us the chance to perform this particular research which is valuable in clinical practice for informed decision making.

